# Effects of Internet Adoption on Depressive Scores and Self-Rated Health in China: Multi-Period Difference-in-Differences Analysis

**DOI:** 10.64898/2026.11.25.26369023

**Authors:** Yu Yun, Yan Dora Zhang

## Abstract

**Background:** Internet use has grown rapidly worldwide over the past decades. Extensive research has explored the health implications of internet use across diverse demographic groups, but most existing studies rely on associational designs. Evidence on the population-level causal effects of internet adoption on both psychological well-being and overall health status remains limited, particularly in settings where individuals adopt the internet across different time points.

**Objective:** This study aimed to investigate the dynamic causal effects of internet adoption on both depressive scores and self-rated health among baseline internet nonusers in China.

**Methods:** This study used longitudinal data from 4 waves of the China Family Panel Studies spanning 2016-2022. Participants who were internet nonusers at the 2016 baseline and provided complete responses across all 4 waves were included, whereas those who ever discontinued internet use after initial adoption were excluded. Internet use was assessed via self-reported questionnaires, depressive scores were evaluated using the Center for Epidemiologic Studies Depression Scale Short Form (CES-D 8), and overall health status was measured using self-rated health on a 5-point scale. A Difference-in-Differences framework with multiple time periods was applied to estimate the causal effects of internet adoption on both outcomes under the conditional parallel trends assumption. Robustness checks and mechanism analyses were further conducted to verify result validity and explore underlying pathways.

**Results:** A total of 4026 participants were included in the analysis. Internet adoption reduced depressive scores on the CES-D 8 scale under a simple weighted average of all group-time average treatment effects (overall ATT = -0.302, SE = 0.156, *P* < .10), whereas event-study aggregation across post-adoption periods revealed a statistically significant cumulative reduction that intensified with longer exposure durations (overall dynamic ATT = -0.354, SE = 0.179, *P* < .05). In contrast, the average treatment effect on self-rated health was not statistically different from 0 (overall ATT = 0.039, SE = 0.046). Mechanism analyses suggested that internet adoption may have lowered depressive scores through expanded engagement in media entertainment, more frequent physical exercise, and higher total family expenditure, whereas the physiological gains of physical exercise may have been offset by prolonged sedentary behavior and health-compromising habits such as smoking. Robustness checks confirmed that these findings were not driven by random chance or sample selection bias.

**Conclusions:** This study demonstrated that among baseline internet nonusers in China, internet adoption generated a cumulative reduction in depressive scores relative to what would have happened in the absence of internet adoption, yet had no statistically significant effect on self-rated health. Future public health initiatives should prioritize inclusive digital infrastructure expansion and age-friendly adaptations alongside fostering digital health literacy, curating health-oriented online content, and implementing targeted measures to mitigate the physiological risks of sedentary behavior and health-compromising habits.

## Introduction

Over the past decades, information and communication technologies have expanded rapidly worldwide [1,2]. The number of internet users has grown from approximately 1 billion in the early 2000s to nearly three-quarters of the global population (about 6 billion people) by 2025 [3]. Digital technologies are now deeply embedded in daily life and have reshaped communication, consumption, and entertainment. As one of the world’s largest digital societies, China has experienced particularly rapid digital transformation, driven by the government’s sustained commitment to expanding internet access. Supported by national policies such as the “Internet Plus” and “Digital Village” initiatives, the internet penetration rate in China surpassed 80% by the end of 2025 [4]. In this context, China provides an important empirical setting for examining the population-level health impacts of internet expansion.

The public health implications of internet adoption have received increasing attention across diverse demographic and health domains. A substantial body of literature has investigated the multifaceted health impacts of internet use among middle-aged and older adults, documenting associations with both mental health [5–8], such as reduced depression levels and better subjective well-being, and physical health outcomes [6,9]. Concurrently, another strand of research has focused on younger populations, exploring how digital engagement and internet use trajectories affect adolescents’ mental health [10–12]. Additionally, research has examined how digital transformation reshapes healthcare-seeking behaviors and service delivery [13–16]. Taken together, these studies underscore the public health relevance of understanding how internet adoption affects population health.

Despite these extensive empirical contributions, establishing a robust causal link between internet use and population health remains challenging. Methodologically, many studies rely either on longitudinal trajectory modeling with internet usage time [7,11] or on associational designs [5,6], often combined with linear mixed models and logistic regressions. Although informative for describing associations, these approaches have limitations in identifying causal effects. In addition, self-reported internet usage time is susceptible to substantial measurement error in longitudinal studies. Even among studies attempting causal inference under multi-period settings, many are constrained by a canonical difference-in-differences (DiD) design [8] or standard two-way fixed effects (TWFE) specifications [12,13,15]. A canonical DiD setup is restricted to 2 periods and 2 groups, whereas real-world digital expansion unfolds across multiple time periods. Meanwhile, TWFE regressions can yield biased estimates in staggered adoption settings with heterogeneous treatment effects due to negative weighting and contaminated comparisons between newly treated and already-treated units [17,18]. Furthermore, multi-period internet adoption raises critical questions regarding whether treatment effects evolve over exposure duration and vary across adoption cohorts. However, standard TWFE regressions compress these potentially dynamic effects into a single average estimate, failing to capture variation across time and cohorts. From a scope perspective, existing studies predominantly focus on isolated age groups and a single health domain, particularly mental health. Consequently, a critical empirical gap remains regarding the population-level causal effects of internet adoption on both mental and physical health outcomes.

To address these gaps, this study employs a quasi-experimental design using 4 waves of longitudinal data from the nationally representative China Family Panel Studies (CFPS) spanning 2016-2022. Specifically, we aim to evaluate the dynamic causal effects of internet adoption on both depressive scores and self-rated health among baseline internet nonusers in China.

## Methods

### Data Source

The data used in this study were obtained from the CFPS, a nationally representative longitudinal survey launched by the Institute of Social Science Survey at Peking University [19,20]. Following the 2010 baseline survey of sampled households and all resident family members, follow-up waves have been conducted biennially, with the 2022 wave as the latest available release. Collecting longitudinal information at individual, family, and community levels, the CFPS covers 25 provinces or their administrative equivalents in China, representing approximately 95% of the national population [19,20].

### Study Sample

To mitigate measurement inconsistency bias arising from questionnaire revisions across survey waves, we prioritized waves with consistent questionnaire items for our core measurements. Starting from 2016, the CFPS assessed internet use through separate questions capturing access via mobile devices and computers, differing from the measurement approach in earlier waves. For depressive scores, the Kessler Psychological Distress Scale (K6) was used in 2014, whereas the Center for Epidemiologic Studies Depression Scale Short Form (CES-D 8) was applied from 2016 onward. Self-rated health was consistently assessed on an identical 5-point scale. Accordingly, we selected the 2016, 2018, 2020, and 2022 waves for this study.

### Study Design and Participants

Since the selected data span 4 time periods and individuals initiated internet access at different time points, a staggered DiD framework with multiple time periods was appropriate for this study [21]. To evaluate the causal effects of internet adoption on both health outcomes, we employed a quasi-experimental design with internet adoption as the treatment. Under the staggered treatment adoption framework, no participant is treated in the initial period, and once treated, participants remain treated throughout subsequent periods. Therefore, individuals who were already using the internet at the 2016 baseline were excluded.

In 2016, the sample included 19,888 participants who completed the self-report questionnaire and were not using the internet. By 2022, only 6293 individuals who participated in all 4 waves remained. Individuals with missing data on key analytical variables were excluded, resulting in a refined sample of 4854 participants. Moreover, to satisfy the staggered treatment adoption assumption, we identified 828 individuals who ever discontinued internet use after initial adoption during the study period and further excluded these “treatment quitters”. Finally, a balanced panel of 4026 participants was constructed for the main analysis. Figure 1 illustrates the participant selection process of this study.

**Figure 1.**
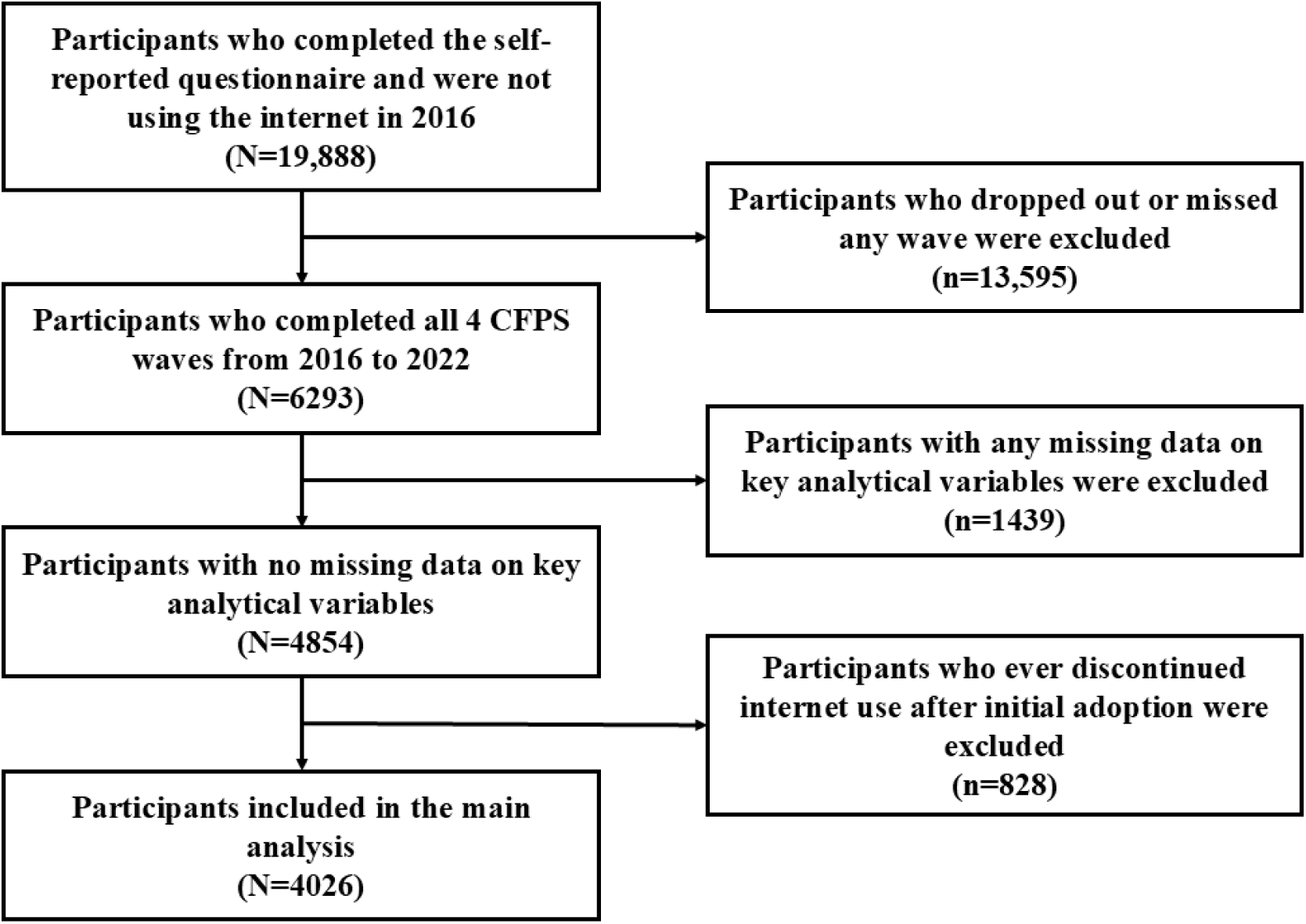
Flow chart of participant selection in this study. CFPS: China Family Panel Studies

### Measurements

#### Internet Use

For all 4 waves, internet use was assessed via two questions: (1) “Do you use mobile devices, for example, mobile phone or tablet PC, to access the internet?” and (2) “Do you use a computer to access the internet?” Responses to both items were recorded as “Yes” or “No.” Internet use was defined as a binary variable, coded as 1 if the participant accessed the internet using either a mobile device or a computer, and 0 otherwise.

#### Depressive Scores

Depressive scores were measured using the CES-D 8, an 8-item self-report scale assessing recent behaviors and emotions [22]. Participants reported the frequency of experiencing the following eight emotional and behavioral states over the past week: (1) “I felt down,” (2) “I found it difficult to do anything,” (3) “I could not sleep well,” (4) “I felt happy,” (5) “I felt lonely,” (6) “I enjoyed life,” (7) “I felt sad,” and (8) “I felt that life could not move on.” Response options were on a 4-point scale: “Hardly ever” (less than 1 day), “Sometimes” (1-2 days), “Often” (3-4 days), and “Most of the time” (5-7 days). Each item was scored from 1 to 4, except for items (4) and (6), which were reverse-coded from 4 to 1 to account for positive emotions. Total scores ranged from 8 to 32 points, with higher scores indicating more severe depressive symptoms.

#### Self-Rated Health

Self-rated health is a widely used valid measure of an individual’s overall health status [23]. It was assessed using a single item: “How would you rate your health status?” Response options included “Excellent,” “Very good,” “Good,” “Fair,” and “Poor.” Responses were coded on a scale of 1 to 5, with higher scores representing poorer self-rated health.

#### Covariates and Behavioral Variables

Covariates were selected based on relevant literature and measurement consistency across survey waves for both children and adults [7,11,13]. We grouped control variables into 3 categories: individual demographic characteristics, individual economic and welfare status, and household-level attributes. Individual demographic variables comprised gender (male or female), age, marital status, and educational attainment. Individual economic and welfare variables included residential area (urban or rural), employment status, and medical insurance coverage. Household-level variables included family size, the natural logarithm of per capita net family income, and the natural logarithm of total family expenditure. Controlling for these baseline covariates helped mitigate confounding bias. Definitions and descriptive statistics for the covariates are listed in Table 1.

**Table 1.**
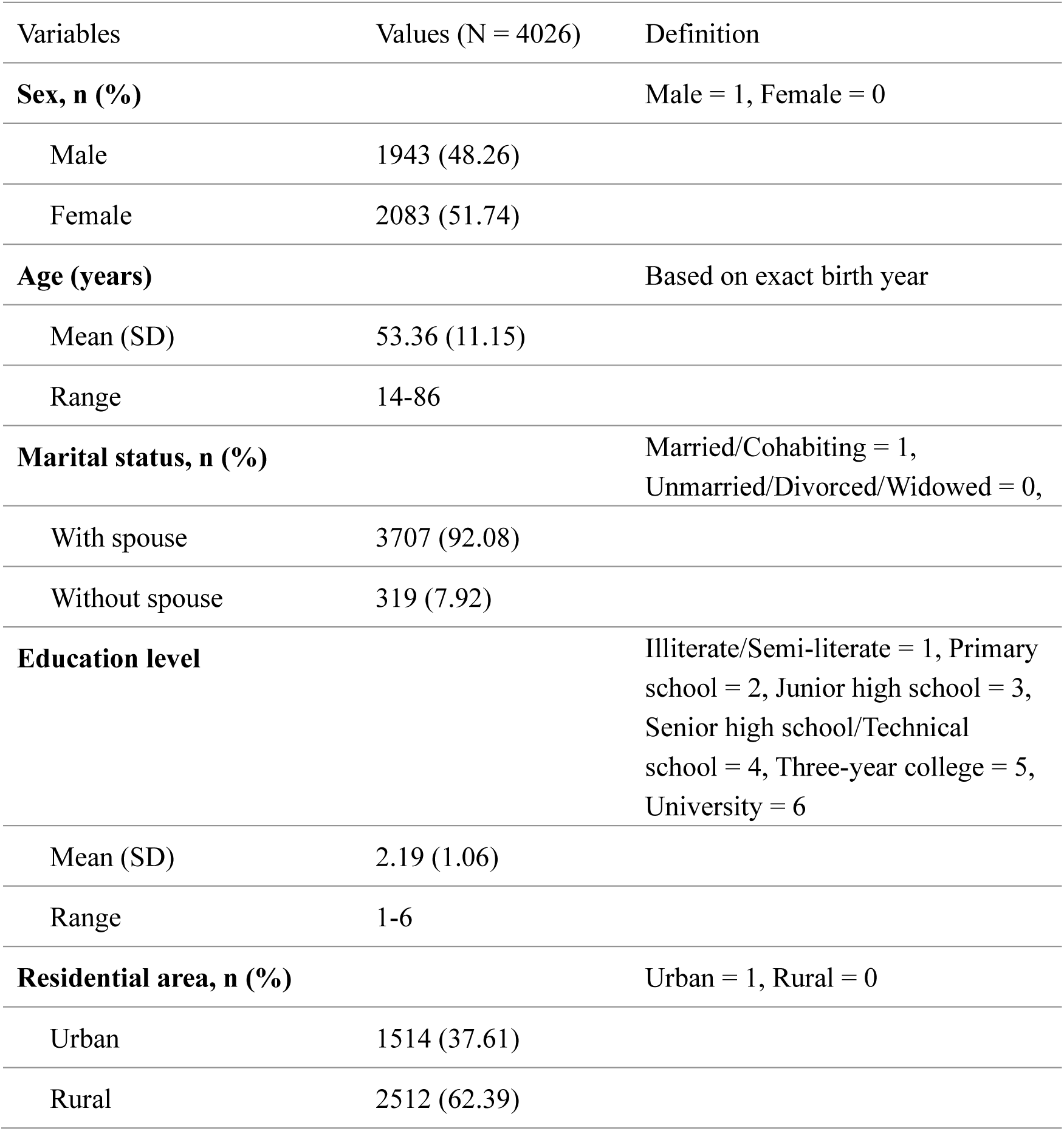
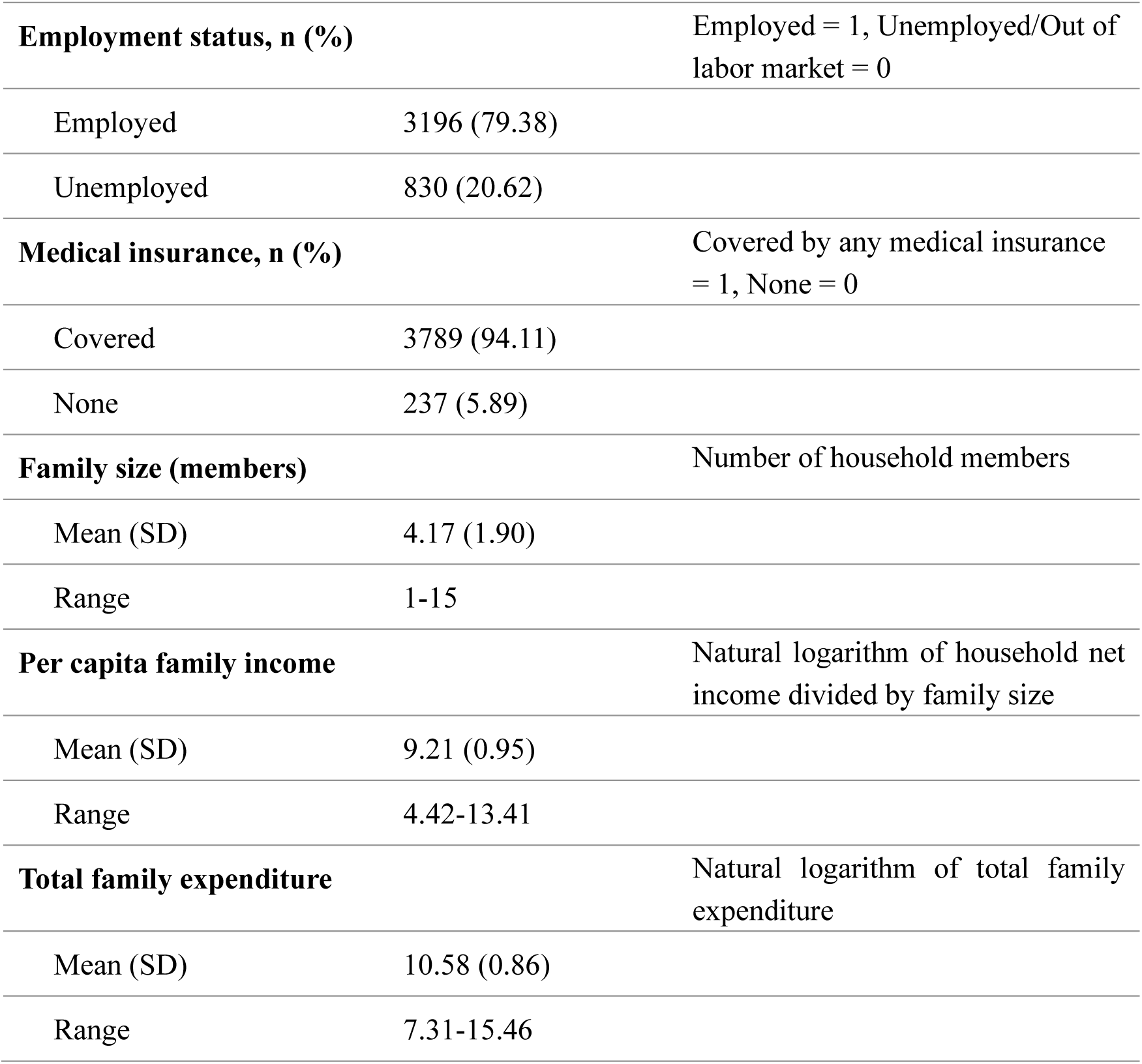
Covariate definitions and baseline population characteristics in 2016.

| Variables | Values (N = 4026) | Definition |
|---|---|---|
| **Sex, n (%)** |  | Male = 1, Female = 0 |
| Male | 1943 (48.26) |  |
| Female | 2083 (51.74) |  |
| **Age (years)** |  | Based on exact birth year |
| Mean (SD) | 53.36 (11.15) |  |
| Range | 14-86 |  |
| **Marital status, n (%)** |  | Married/Cohabiting = 1, Unmarried/Divorced/Widowed = 0, |
| With spouse | 3707 (92.08) |  |
| Without spouse | 319 (7.92) |  |
| **Education level** |  | Illiterate/Semi-literate = 1, Primary school = 2, Junior high school = 3, Senior high school/Technical school = 4, Three-year college = 5, University = 6 |
| Mean (SD) | 2.19 (1.06) |  |
| Range | 1-6 |  |
| **Residential area, n (%)** |  | Urban = 1, Rural = 0 |
| Urban | 1514 (37.61) |  |
| Rural | 2512 (62.39) |  |
| **Employment status, n (%)** |  | Employed = 1, Unemployed/Out of labor market = 0 |
| Employed | 3196 (79.38) |  |
| Unemployed | 830 (20.62) |  |
| **Medical insurance, n (%)** |  | Covered by any medical insurance = 1, None = 0 |
| Covered | 3789 (94.11) |  |
| None | 237 (5.89) |  |
| **Family size (members)** |  | Number of household members |
| Mean (SD) | 4.17 (1.90) |  |
| Range | 1-15 |  |
| **Per capita family income** |  | Natural logarithm of household net income divided by family size |
| Mean (SD) | 9.21 (0.95) |  |
| Range | 4.42-13.41 |  |
| **Total family expenditure** |  | Natural logarithm of total family expenditure |
| Mean (SD) | 10.58 (0.86) |  |
| Range | 7.31-15.46 |  |

Additionally, we identified several individual behavioral variables to explore potential mechanisms, including smoking, alcohol consumption, physical exercise frequency, and media entertainment time. Smoking was defined as a binary indicator for having smoked in the past month. Alcohol consumption was defined as a binary variable indicating whether the individual drank alcohol at least 3 times per week. Physical exercise frequency was assessed via the question: “Including physical education, in the past 12 months, how often did you participate in physical fitness and leisure activities?” Since the response format changed across waves from numerical counts to categorical options, we standardized the variable into 4 ordinal categories to ensure measurement consistency: 0 for no participation, 1 for exercising 1-2 times per week, 2 for 3-6 times per week, and 3 for at least 7 times per week. Media entertainment time was assessed using the question: “In general, how long do you usually spend watching TV, movies, or videos every week?” and was recorded in hours per week. Descriptive statistics for these behavioral variables across waves are provided in Table 2.

**Table 2.** Core measurements and behavioral variables across waves (N = 4026).

| Variables | Values in 2016 | Values in 2018 | Values in 2020 | Values in 2022 |
|---|---|---|---|---|
| **Internet use, n (%)** |  |  |  |  |
| Yes | 0 (0) | 740 (18.38) | 1289 (32.02) | 1917 (47.62) |
| No | 4026 (100.00) | 3286 (81.62) | 2737 (67.98) | 2109 (52.38) |
| **Depressive scores (CES-D 8)** |  |  |  |  |
| Mean (SD) | 13.33 (4.24) | 13.63 (4.04) | 13.83 (4.34) | 14.18 (4.40) |
| Range | 8-32 | 8-32 | 8-32 | 8-32 |
| **Self-rated health** |  |  |  |  |
| Mean (SD) | 3.20 (1.26) | 3.20 (1.26) | 3.14 (1.30) | 3.17 (1.29) |
| Range | 1-5 | 1-5 | 1-5 | 1-5 |
| **Smoking, n (%)** |  |  |  |  |
| Yes | 1158 (28.76) | 1199 (29.78) | 1124 (27.92) | 1100 (27.32) |
| No | 2868 (71.24) | 2827 (70.22) | 2902 (72.08) | 2926 (72.68) |
| **Alcohol consumption, n (%)** |  |  |  |  |
| Yes | 668 (16.59) | 665 (16.52) | 578 (14.36) | 567 (14.08) |
| No | 3358 (83.41) | 3361 (83.48) | 3448 (85.64) | 3459 (85.92) |
| **Physical exercise frequency** |  |  |  |  |
| Mean (SD) | 0.97 (1.30) | 1.18 (1.36) | 0.64 (1.15) | 0.74 (1.21) |
| Range | 0-3 | 0-3 | 0-3 | 0-3 |
| **Media entertainment time (hours/week)** |  |  |  |  |
| Mean (SD) | 12.67 (10.36) | 11.11 (10.49) | 9.82 (10.51) | 8.85 (10.97) |
| Range | 0-105 | 0-144 | 0-150 | 0-150 |

### Statistical Analysis

#### Two-Way Fixed Effects Regression

We first employed a standard TWFE linear regression to estimate the causal effects for the panel data as an initial benchmark [24]. Since the primary approach applied in this study, DiD with multiple time periods, relies on pre-treatment covariates to satisfy the conditional parallel trends assumption, we incorporated interaction terms between covariates measured at the 2016 baseline and a standardized survey wave index into the TWFE model. This specification not only mitigated potential endogeneity arising from time-varying covariates but also ensured that any discrepancy between TWFE and multi-period DiD estimates could be attributed to methodological differences rather than variations in covariates. The TWFE regression was performed using the “fixest” package in R.

The baseline TWFE model is specified as follows:

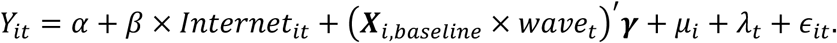

Here, *Y_it_* denotes the depressive score or self-rated health of individual *i* at time period *t*. *Internet_it_* is the indicator for internet use, and *β* is the parameter capturing the average treatment effect of internet use. ***X****_i_*_,*baseline*_ includes the covariates for individual *i* measured at the 2016 baseline, and 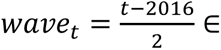 {0,1,2,3} denotes the standardized survey wave index corresponding to the 2016, 2018, 2020, and 2022 CFPS waves. *μ_i_* and *λ_t_* represent individual and time fixed effects, respectively, and *ε_it_* is the error term.

However, recent econometric studies have demonstrated that standard TWFE estimates can be biased and unreliable for causal inference in settings with staggered treatment adoption due to contaminated comparisons [17,18]. To diagnose the reliability of the TWFE regression and assess the proportion of problematic comparisons, we conducted a Goodman-Bacon decomposition using the R package “bacondecomp” [25]. These methodological concerns directly motivated our application of the staggered DiD framework, which is specifically designed to overcome the limitations of conventional TWFE regressions.

#### Difference-in-Differences with Multiple Time Periods

To estimate the causal effects under a staggered treatment adoption setting across the 4 CFPS waves, we applied DiD with multiple time periods. This framework estimates the group-time average treatment effect for cohort *g* at time period *t*, denoted by:

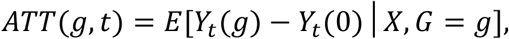

where cohort *g* is defined by the time period when individuals are first treated [21]. *Y_t_*(*g*) is the potential outcome in time period *t* if individuals become treated in period *g* and *Y_t_*(0) denotes the untreated potential outcome. Counterfactuals are constructed using either never-treated or not-yet-treated individuals, thereby avoiding the use of already-treated individuals as controls. Unlike the unconditional parallel trends assumption required by canonical DiD and TWFE setups, this framework relies on the conditional parallel trends assumption, which holds after conditioning on pre-treatment covariates ***X***. This assumption is substantially more practical and well-suited to our empirical setting, because it appropriately accommodates covariate-specific trends in outcomes over time and acknowledges that the covariate distributions may differ across adoption cohorts. The group-time average treatment effect naturally reveals how treatment effects vary across cohorts and time periods. Moreover, the group-time average treatment effects can be aggregated into composite causal parameters to address specific questions regarding treatment effect heterogeneity.

DiD with multiple time periods was performed using the “did” package in R. To satisfy the conditional parallel trends assumption, we incorporated the pre-treatment covariates ***X****_i_*_,*baseline*_ measured at the 2016 baseline into the conditioning specification. Since around half of the participants remained untreated in the final wave, we selected never-treated individuals as the control group. Because internet adoption was gradual and individual-specific rather than tied to a clearly scheduled intervention, treatment anticipation was considered unlikely, and the anticipation period was set to 0. With these assumptions satisfied and the never-treated units serving as controls, the group-time average treatment effect for cohort *g* at time period *t* is identified by:

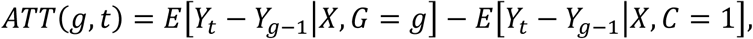

where *Y_t_* and *Y_g_*_−1_ denote the outcomes at time *t* and the preceding pre-treatment period *g* − 1, respectively, and *C* is an indicator for whether an individual is in the never-treated group. Given our reliance on the conditional parallel trends assumption, we employed the doubly robust estimation method [21]. To comprehensively evaluate treatment effect heterogeneity, we examined several aggregated summary measures of group-time average treatment effects, including aggregation by length of exposure, by adoption cohort, and across time periods.

#### Robustness Check

To ensure the reliability of our empirical findings, we conducted 2 robustness checks to rule out the possibility that the observed effects were driven by random chance or unobserved confounders and to address sample selection concerns.

First, we conducted a placebo test by randomly assigning both treatment timing and treatment status across individuals [13]. We repeated this Monte Carlo simulation 500 times, re-estimating the treatment effects in each iteration to generate an empirical null distribution of placebo estimates tightly centered around 0. Observed estimates falling far outside this null distribution indicate genuine causal effects rather than artifacts of random chance.

Second, as detailed in the sample selection procedure, individuals who ever discontinued internet use after initial adoption (treatment quitters) were excluded from the primary sample to satisfy the staggered treatment adoption assumption. To ensure that this restriction did not introduce selection bias or invalidate our conclusions, we reintroduced these treatment quitters into the full sample and employed an alternative estimator for event-study effects. This alternative framework allows individuals to move in and out of treatment status over time, thereby enabling us to verify that our findings remain robust to behavioral fluctuations in internet use [26]. The estimator was obtained using the “DIDmultiplegtDYN” package in R [27]. Since this method relies on a fast fixed-effects estimation approach, we incorporated the same interaction terms between baseline covariates and the survey wave index (***X****_i_*_,*baseline*_ × *wave_t_*) as in the TWFE model. This specification maintained methodological consistency across models and reduced bias arising from differential time trends associated with covariates.

### Ethical Considerations

The CFPS received ethical review approval from the Peking University Biomedical Ethics Review Committee (approval number: IRB00001052-14010), and informed consent was obtained from all eligible participants. Since this study was based solely on a secondary analysis of anonymized CFPS data, no additional ethical approval was required after retrieving the data from the official repository.

## Results

### Descriptive Statistics

Table 1 summarizes the baseline characteristics of the analytical sample (N = 4026) in 2016. Overall, the sample featured a relatively balanced sex distribution (2083 female participants, 51.74%) and a wide age span (mean = 53.36; SD = 11.15; range 14-86 years), capturing a broad age distribution among baseline internet nonusers that leaned toward middle-aged and older adults. Most participants were married or cohabiting (n **=** 3707, 92.08%). The mean education level was 2.19 (SD = 1.06; range 1-6), reflecting overall educational attainment predominantly below the senior high school level. In terms of socioeconomic indicators, 2512 respondents (62.39%) resided in rural areas, 3196 (79.38%) were employed, and most respondents (n = 3789, 94.11%) were covered by medical insurance. At the household level, the mean family size was 4.17 members (SD = 1.90). The means of log-transformed per capita family income and log-transformed total family expenditure were 9.21 (SD = 0.95) and 10.58 (SD = 0.86), respectively.

Table 2 outlines the longitudinal trends of health outcomes, internet adoption, and behavioral variables across the 4 waves within the study sample. Internet adoption expanded steadily during the study period, with adoption rates increasing by roughly 15 percentage points per wave (n = 740, 18.38%, in 2018; n = 1289, 32.02%, in 2020; n = 1917, 47.62%, in 2022). By 2022, nearly half of the sample had adopted the internet, whereas the other half remained in the never-treated group, providing a sufficiently large control group for causal identification. In terms of health outcomes, mean CES-D 8 scores increased incrementally from 13.33 (SD = 4.24) in 2016 to 14.18 (SD = 4.40) in 2022, suggesting an overall worsening trend in depressive symptoms over time. Conversely, self-rated health remained relatively stable across waves. Regarding behavioral variables, the proportions of smoking and alcohol consumption showed slight downward trends. Physical exercise frequency fluctuated modestly, peaking in 2018. Mean weekly media entertainment time decreased progressively across waves, accompanied by an expanding range and increasing SD. Although these descriptive statistics illustrate unconditional general trends, quasi-experimental estimation is essential to uncover the causal effects of internet adoption on these outcomes.

### TWFE Estimates and Model Diagnostics

Table 3 presents the TWFE regression results. Model 1 refers to the specification without control variables, and Model 2 incorporates the full baseline covariate interaction structure. The coefficients for internet use are not statistically significant for either depressive scores or self-rated health in either specification. Specifically, Model 2 yielded point estimates of -0.132 (SE = 0.099) for depressive scores and -0.007 (SE = 0.029) for self-rated health. Although not statistically significant, the negative and sizable point estimates for depressive scores suggest a potentially inverse relationship between internet adoption and depressive scores, whereas the estimates for self-rated health were close to 0.

**Table 3.**
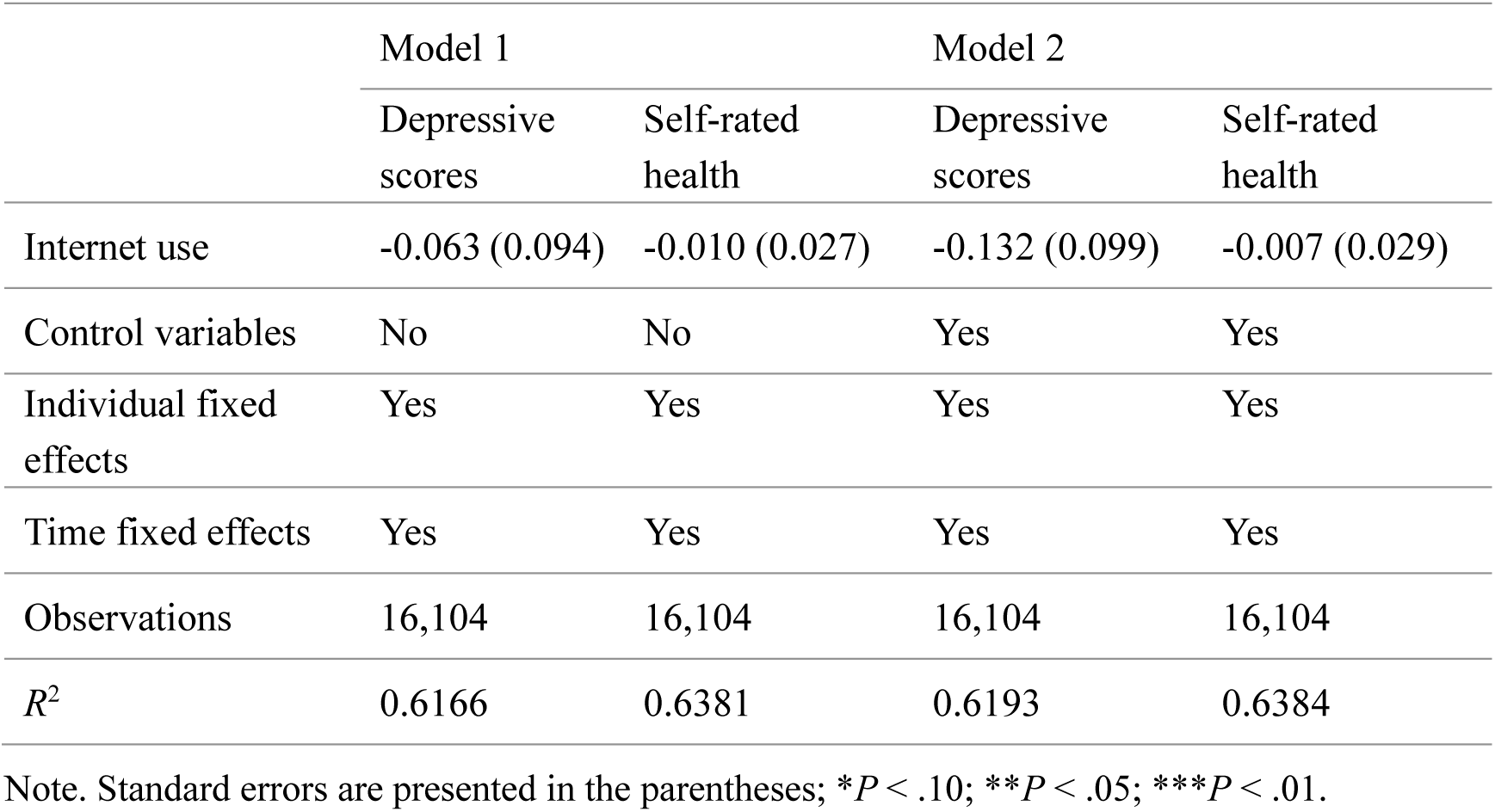
TWFE regression results.

Table 4 reports the Goodman-Bacon decomposition results for the uncontrolled TWFE regression. Although comparisons between treated and untreated individuals accounted for 76.4% of the total weight, 23.6% derived from timing-based comparisons between different internet adoption cohorts (11.6% from earlier-treated versus later-treated and 12.0% from later-treated versus earlier-treated individuals). Under staggered treatment adoption, using already-treated units as control groups for newly treated cohorts introduces contaminated comparisons. Since these control groups are themselves already treated, their outcomes reflect ongoing treatment dynamics rather than serving as valid counterfactuals. In the presence of treatment effect heterogeneity across time or cohorts, including such comparisons can produce substantial weighting bias and attenuate the composite estimate. Therefore, the notable weight allocated to problematic comparisons renders the TWFE estimates unreliable for causal inference. These diagnostic results strongly justify the necessity of transitioning to the staggered DiD framework.

**Table 4.**
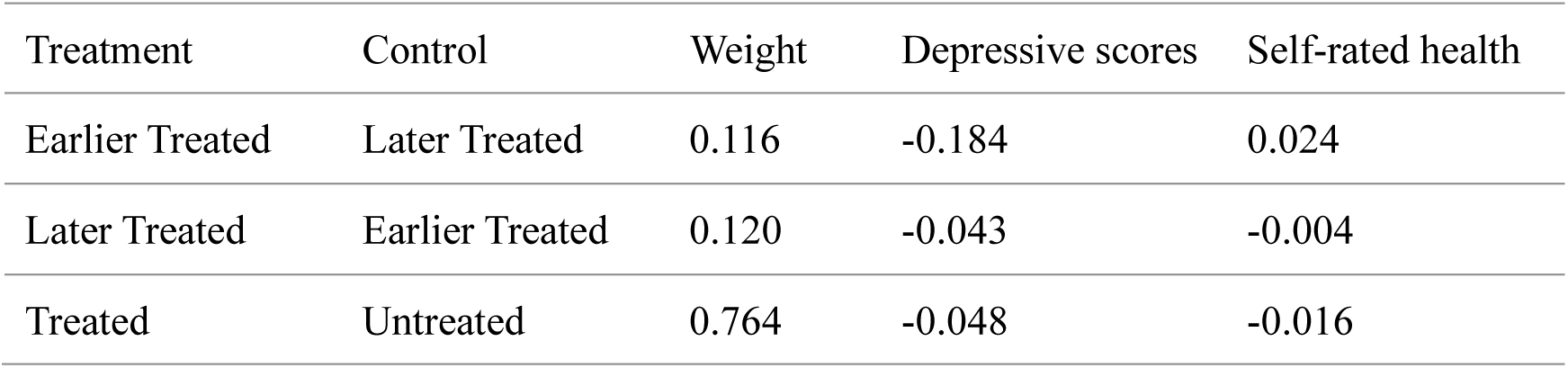
Goodman-Bacon decomposition results for uncontrolled TWFE regression.

### Causal Effect Estimates from DiD with Multiple Time Periods

Table 5 presents 4 types of overall summaries for aggregated causal parameters of internet adoption on depressive scores (top panel) and self-rated health (bottom panel). These summaries include a simple weighted average of all group-time average treatment effects with weights proportional to sample size (Simple), an average effect across different internet adoption cohorts (Group-specific), an average effect across all positive lengths of exposure to internet adoption (Event-study), and an overall effect that averages across each time period (Calendar-time). Models 1-4 progressively incorporate covariates: Model 1 contains no controls; Model 2 includes individual demographic controls; Model 3 further incorporates individual economic and welfare controls; and Model 4 represents the fully controlled model, incorporating all 3 categories of baseline covariates.

**Table 5.** Overall summaries of aggregated group-time average treatment effects.

|  | Model 1 | Model 2 | Model 3 | Model 4 |
|---|---|---|---|---|
| **Depressive scores** |  |  |  |  |
| Simple | -0.126 (0.107) | -0.339** (0.171) | -0.319* (0.167) | -0.302* (0.156) |
| Group-specific | -0.109 (0.103) | -0.267* (0.141) | -0.253* (0.132) | -0.250* (0.136) |
| Event-study | -0.136 (0.121) | -0.389** (0.198) | -0.368** (0.184) | -0.354** (0.179) |
| Calendar-time | -0.113 (0.111) | -0.340* (0.180) | -0.318* (0.171) | -0.285* (0.170) |
| **Self-rated health** |  |  |  |  |
| Simple | 0.0002 (0.030) | 0.048 (0.046) | 0.050 (0.046) | 0.039 (0.046) |
| Group-specific | -0.010 (0.029) | 0.025 (0.037) | 0.028 (0.037) | 0.020 (0.036) |
| Event-study | -0.001 (0.034) | 0.054 (0.050) | 0.058 (0.053) | 0.046 (0.051) |
| Calendar-time | 0.010 (0.031) | 0.050 (0.048) | 0.051 (0.047) | 0.042 (0.044) |

Note. Standard errors are presented in the parentheses; \**P* < .10; \*\**P* < .05; \*\*\**P* < .01.

The results in Table 5 indicate that incorporating pre-treatment covariates is critical in this study, whereas the overall effects across all aggregation dimensions exhibit stability as covariates are progressively included. For depressive scores, overall effects are not statistically significant in the unconditional specification, mirroring the results from the TWFE regression. However, once the covariates are introduced, statistically significant overall effects are observed and remain highly stable from Models 2 to 4 (eg, the overall effects based on event-study aggregation vary between -0.35 and -0.39 and retain statistical significance). In contrast, for self-rated health, the overall effects are not statistically different from 0, although a similar pattern is evident. The transition from the unconditional specification to the controlled models introduces a discernible difference, whereas progressively adding covariates results in a negligible variation (eg, the overall effects based on group-specific aggregation fluctuate narrowly between 0.02 and 0.03). This consistent pattern supports the appropriateness and robustness of our covariate selection. Consequently, subsequent analyses focused on the results from the fully controlled specification.

Figure 2 and Figure 3 present the estimated group-time average treatment effects of internet adoption on depressive scores and self-rated health, respectively, under the unconditional parallel trends assumption (panel A) and the conditional parallel trends assumption (panel B). Detailed effect estimates and standard errors under the conditional specification are available in Table S1 in Multimedia Appendix 1. For depressive scores (Figure 2), the test for the unconditional parallel trends assumption yielded *P* = .098, indicating a violation of the assumption at the 10% significance level without controlling for covariates. After incorporating a comprehensive set of pre-treatment covariates, the conditional parallel trends assumption is satisfied (*P* = .606), supporting the assumption that individuals with comparable baseline characteristics would follow parallel trends in depressive scores in the absence of internet adoption. For self-rated health (Figure 3), both the unconditional and conditional specifications pass the parallel trends test (unconditional, *P* = .995; conditional, *P* = .982). These findings again highlight the necessity of conditioning on pre-treatment covariates and validate the conditional parallel trends assumption in our empirical analysis.

**Figure 2.**
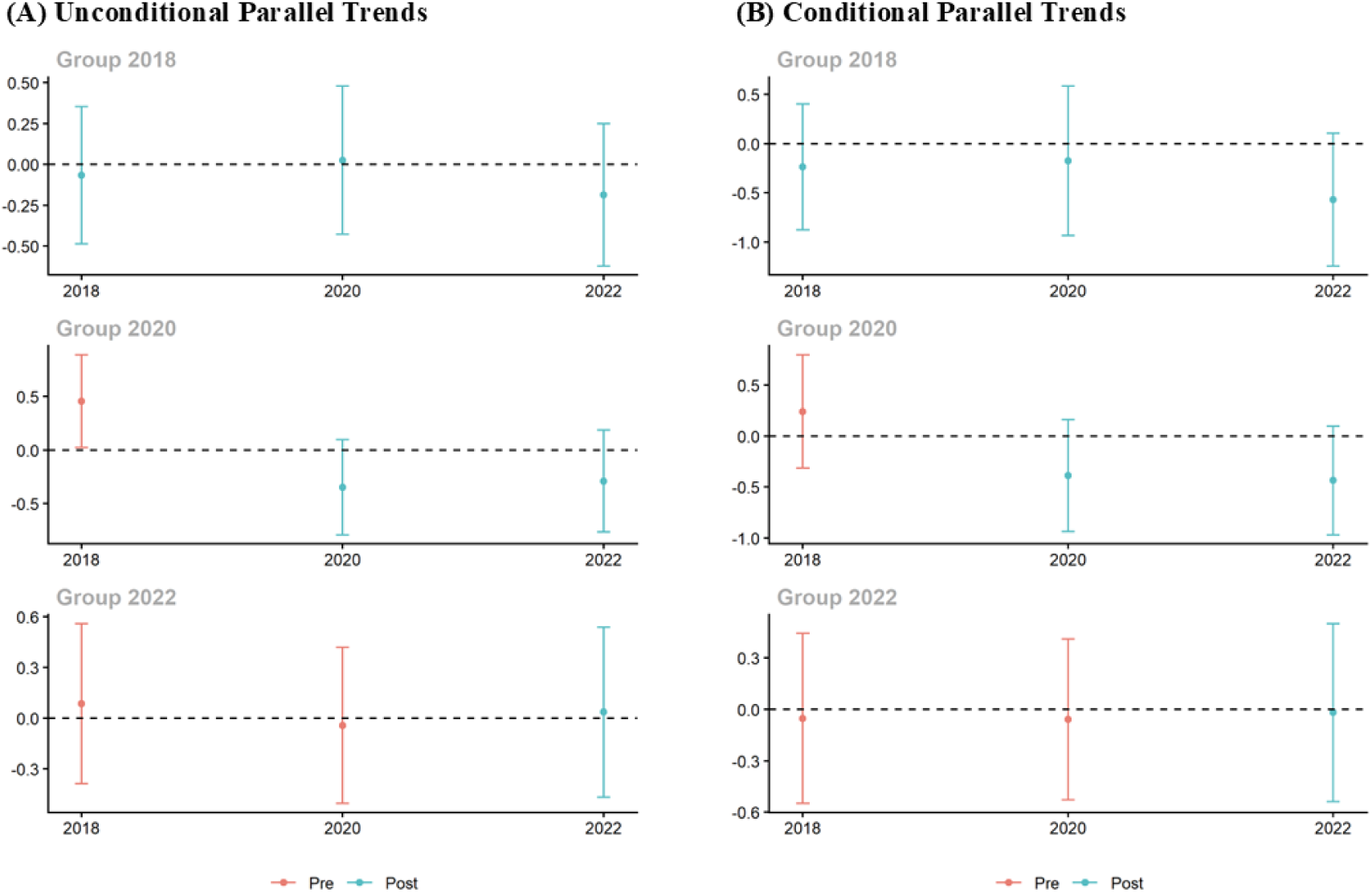
Group-time average treatment effects of internet adoption on depressive scores. **(A)** Estimates under the unconditional parallel trends assumption; **(B)** estimates under the conditional parallel trends assumption. Red dots and error bars represent point estimates and 90% simultaneous confidence intervals for pre-treatment periods, whereas blue dots and error bars represent those for post-treatment periods.

**Figure 3.**
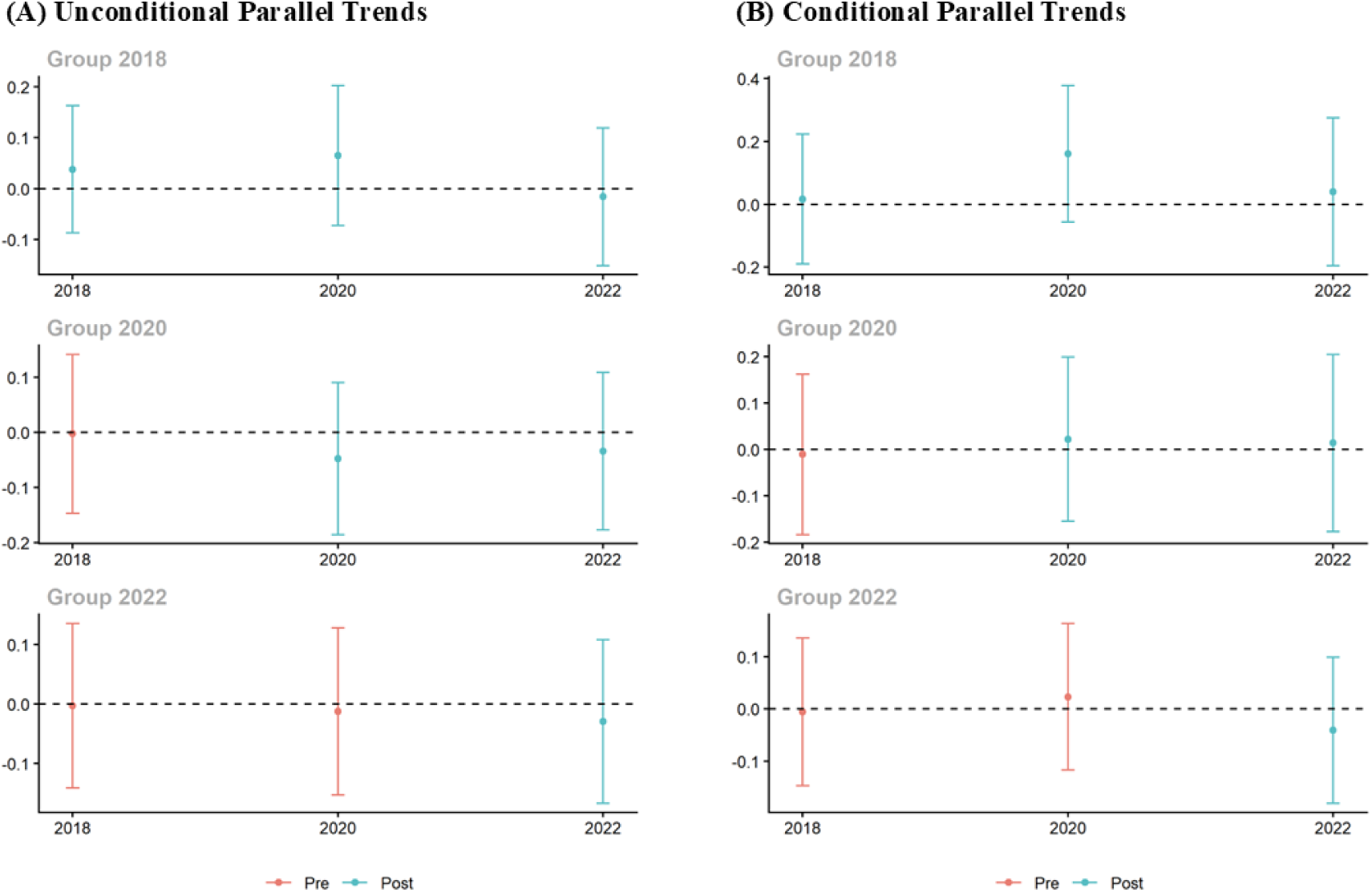
Group-time average treatment effects of internet adoption on self-rated health. **(A)** Estimates under the unconditional parallel trends assumption; **(B)** estimates under the conditional parallel trends assumption. Red dots and error bars represent point estimates and 90% simultaneous confidence intervals for pre-treatment periods, whereas blue dots and error bars represent those for post-treatment periods.

Beyond the overall summaries of aggregated group-time treatment effects, we focused on three specific questions regarding treatment effect heterogeneity: (1) Did average treatment effects differ depending on when individuals first adopted the internet? (2) How did average treatment effects vary with the length of exposure to continuous internet use? (3) What was the cumulative average treatment effect of internet adoption up to a particular time period? To address these questions, we investigated the underlying dynamics of treatment effects across 3 aggregation dimensions.

Figure 4, Figure 5, and Figure 6 present the variations in average treatment effects of internet adoption on depressive scores (panel A) and self-rated health (panel B) based on group-specific, event-study, and calendar-time aggregations, respectively. Detailed effect estimates and standard errors are available in Table S2 in Multimedia Appendix 1. Regarding cohort-level variation, Figure 4 reveals differences in average effects on depressive scores tied to the timing of internet adoption. The effect of internet adoption on depressive scores is negative and larger in magnitude for the earlier adoption cohorts (Cohorts 2018 and 2020), whereas no statistically significant effect is observed for the 2022 cohort. Since group-specific aggregation averages effects across all available post-treatment periods, this pattern suggests a cumulative nature in the effect of internet adoption on depressive scores. This finding is further corroborated by Figure 5, which demonstrates a similar pattern through event-study aggregation. The pre-treatment effects are statistically indistinguishable from 0, reinforcing that the conditional parallel trends assumption is valid. The post-treatment effect of internet adoption on depressive scores is negative, emerging upon initial adoption and increasing in magnitude the longer individuals remain exposed to continuous internet use. In particular, depressive scores are estimated to decrease by 0.209 upon internet adoption (*e* = 0), decrease by 0.286 after 2 years of continuous use (*e* = 2), and by 0.568 with 4 years of continuous use (*e* = 4). Moreover, Figure 6 illustrates the average treatment effects across individuals who had adopted the internet by each calendar time period. Although the estimated reduction gradually increases in magnitude from 0.238 in 2018 to 0.350 in 2020, a statistically significant result is observed by 2022. Despite differences in statistical power primarily driven by increasing adoption rates, this pattern is consistent with the idea that internet adoption had a negative effect on depressive scores that strengthened over time.

**Figure 4.**
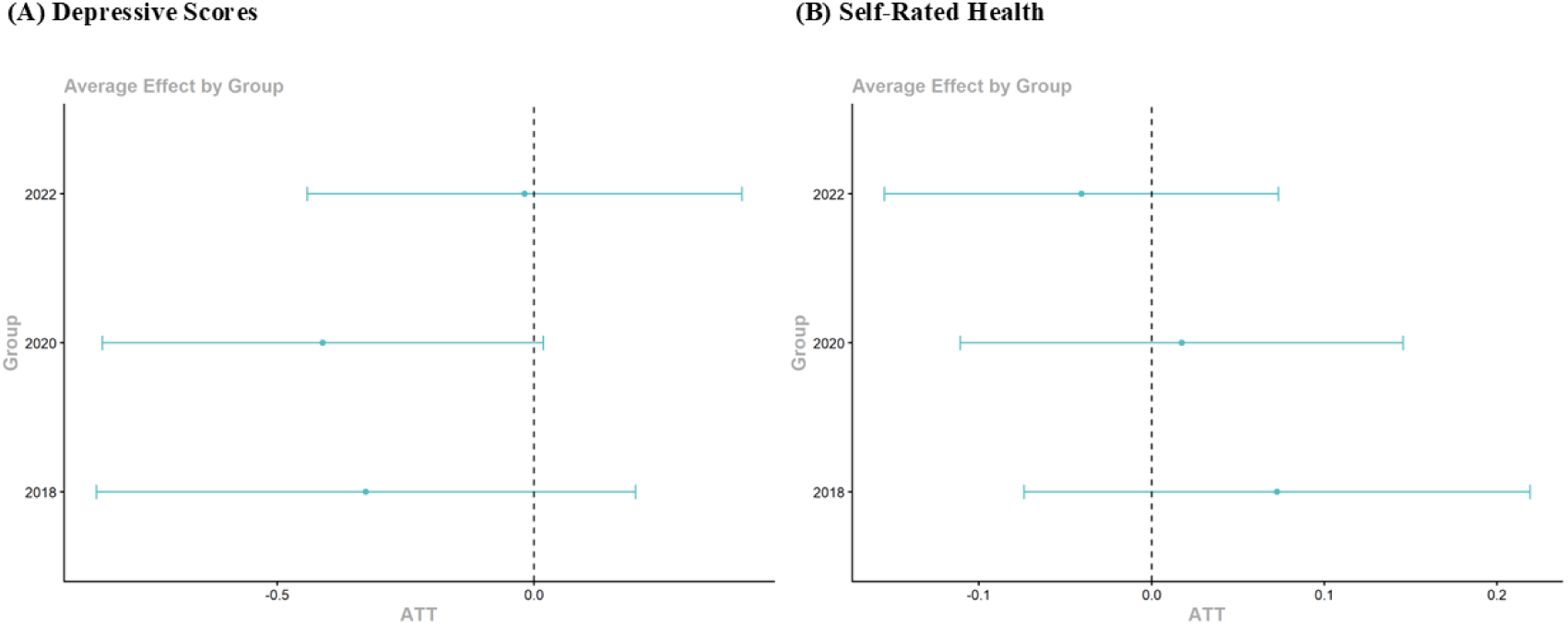
Average treatment effects of internet adoption based on group-specific aggregation. **(A)** Depressive scores; **(B)** Self-rated health. Dots represent point estimates, and error bars indicate 90% simultaneous confidence intervals.

**Figure 5.**
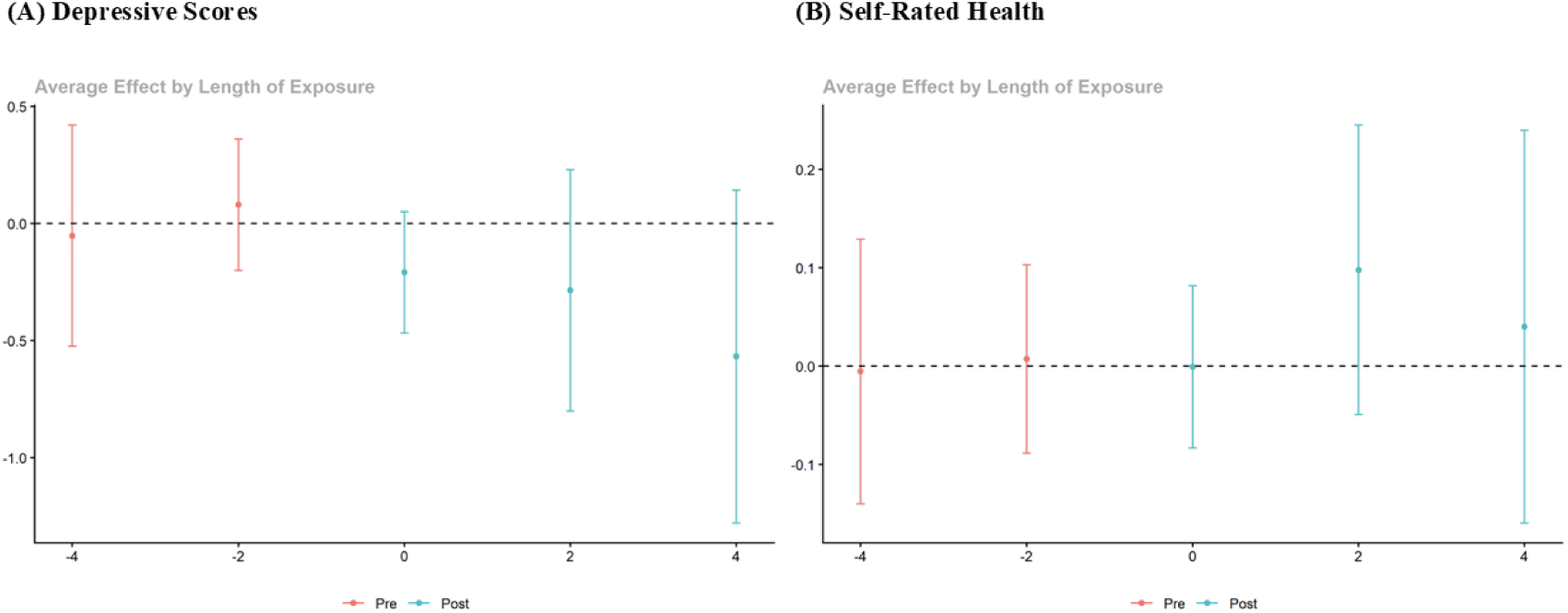
Average treatment effects of internet adoption based on event-study aggregation. **(A)** Depressive scores; **(B)** Self-rated health. Red dots and error bars represent point estimates and 90% simultaneous confidence intervals for pre-treatment periods, whereas blue dots and error bars represent those for post-treatment periods.

**Figure 6.**
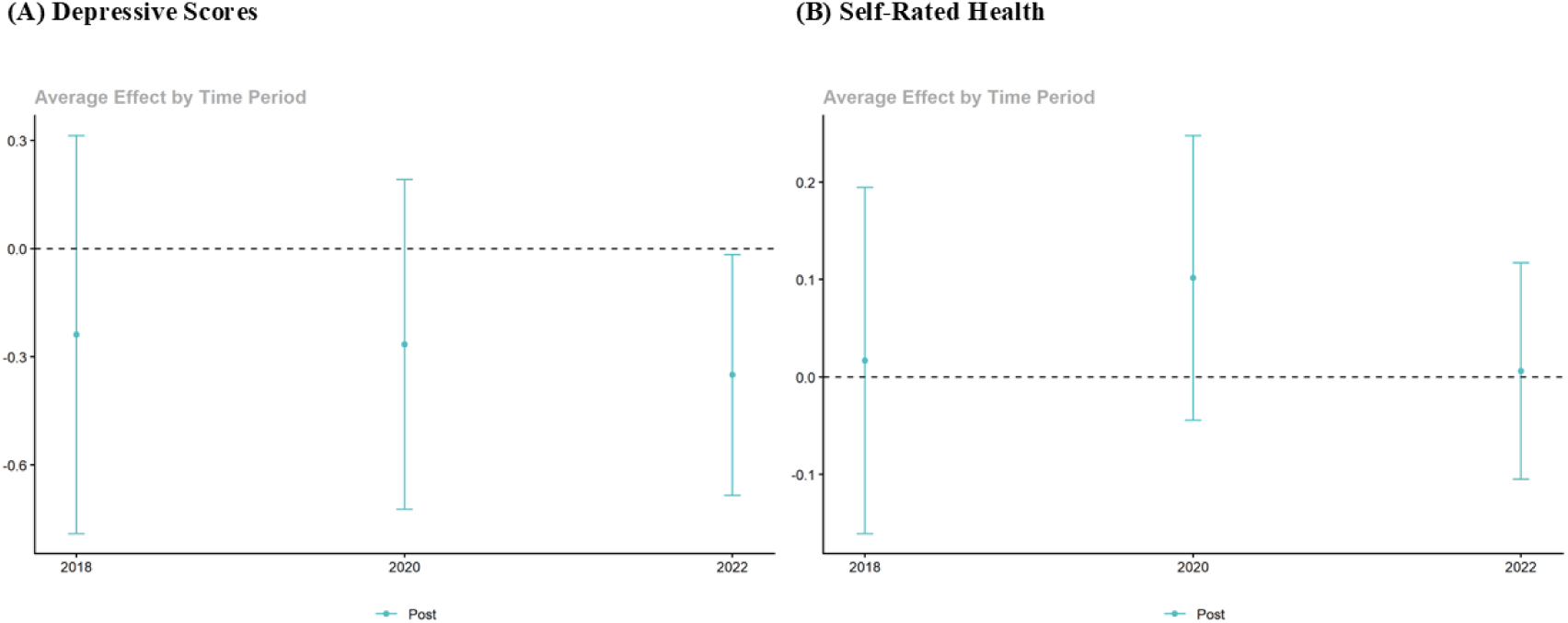
Average treatment effects of internet adoption based on calendar-time aggregation. **(A)** Depressive scores; **(B)** Self-rated health. Dots represent point estimates, and error bars indicate 90% simultaneous confidence intervals.

In contrast, the effect profiles for self-rated health across Figure 4, Figure 5, and Figure 6 show no significant variation before or after internet adoption, and the aggregated effects of internet adoption on self-rated health are not statistically different from 0. These results for self-rated health align with the overall summaries in Table 5. The divergence observed in the aggregated treatment effects between the 2 health outcomes reveals an asymmetric health return pattern. Specifically, internet adoption produced a negative and cumulative effect on depressive scores relative to what would have happened in the absence of internet adoption, whereas it had no statistically significant effect on self-rated health.

### Robustness Analysis

Figure 7 and Figure 8 present the placebo test results for depressive scores and self-rated health, respectively, across all 4 aggregation dimensions (Simple, Group-specific, Event-study, and Calendar-time). As shown in Figure 7, the actual ATTs lie in the extreme tails of the simulated distributions, indicating that random permutations of treatment assignment rarely reproduce the observed effects by chance. This confirms that the negative effect of internet adoption on depressive scores is genuine rather than an artifact of random chance. Conversely, for self-rated health (Figure 8), the actual ATT estimates are centered within the placebo distributions, yielding high empirical *P* values across all 4 aggregation dimensions. These results indicate that the estimated effects on self-rated health are statistically indistinguishable from those generated under random assignment, confirming a null effect. Collectively, these placebo test results supported the robustness of our findings from the multi-period DiD analysis.

**Figure 7.**
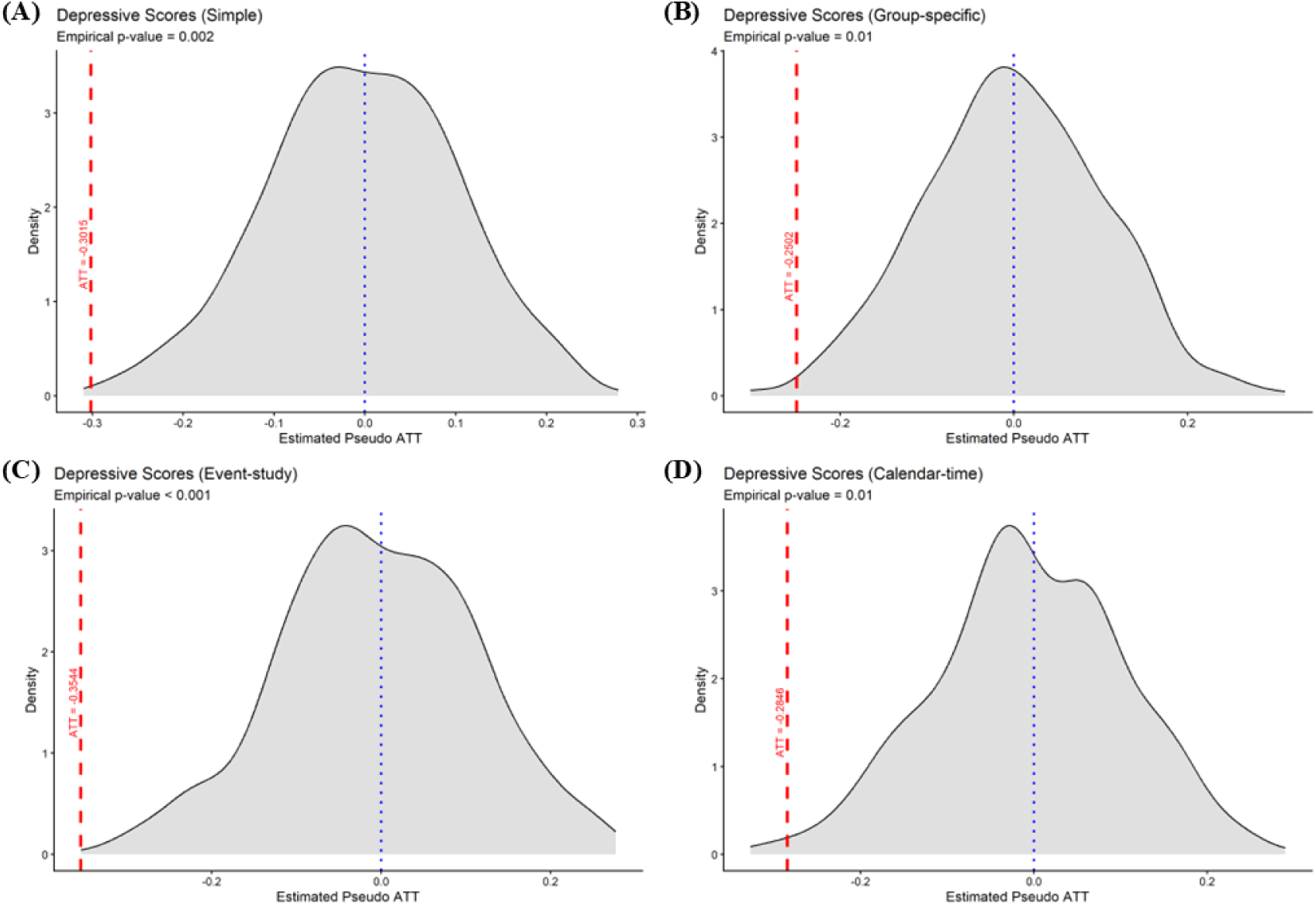
Placebo test results for overall summaries of aggregated effects of internet adoption on depressive scores. **(A)** Simple aggregation; **(B)** group-specific aggregation; **(C)** event-study aggregation; **(D)** calendar-time aggregation. Shaded density curves show the empirical distributions of estimated pseudo-ATTs across 500 random permutations. Vertical red dashed lines denote the actual overall summary of aggregated group-time average treatment effects reported in Table 5. Empirical *P* values are obtained from the permutation tests under the null hypothesis of no effect.

**Figure 8.**
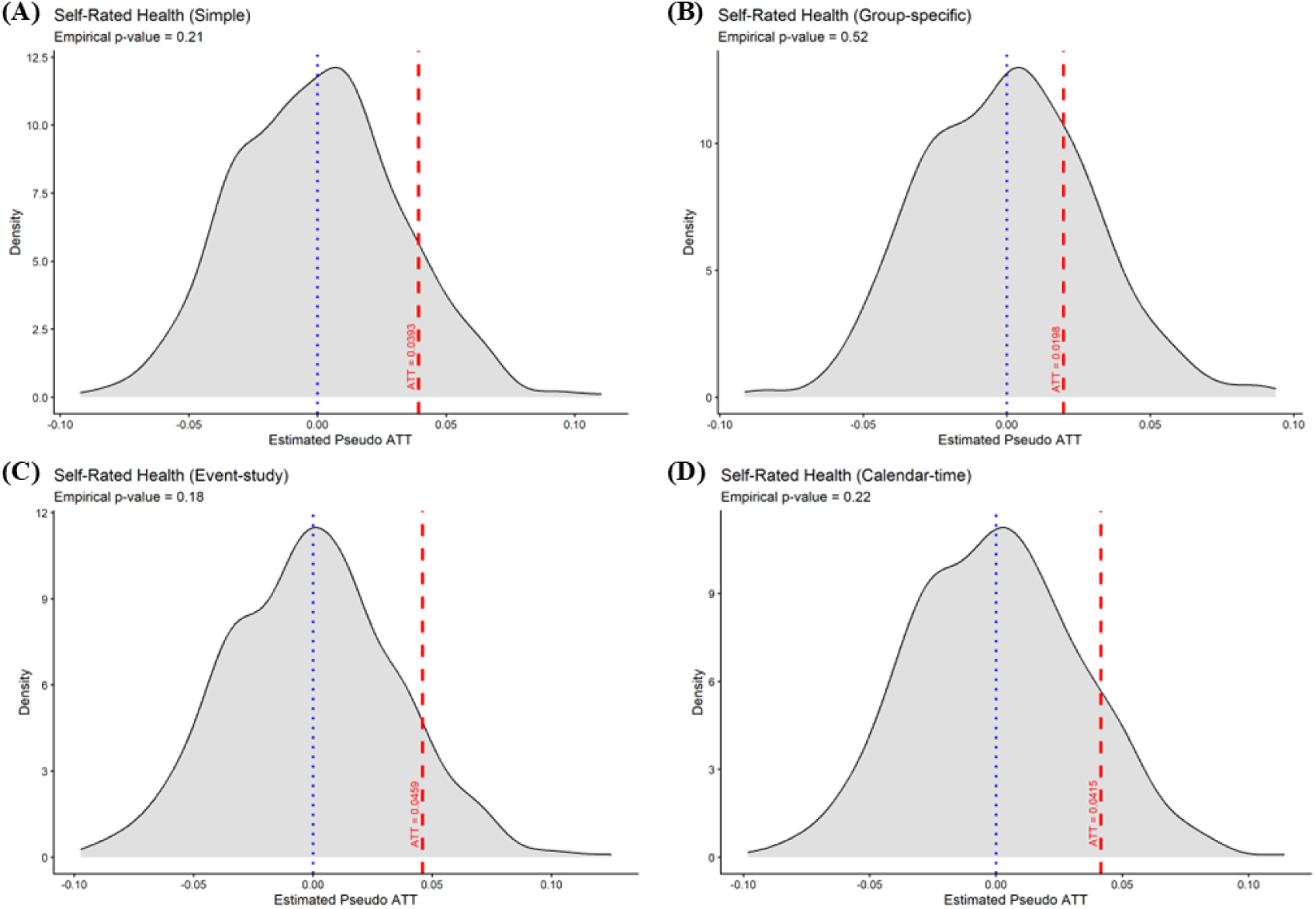
Placebo test results for overall summaries of aggregated effects of internet adoption on self-rated health. **(A)** Simple aggregation; **(B)** group-specific aggregation; **(C)** event-study aggregation; **(D)** calendar-time aggregation. Shaded density curves show the empirical distributions of estimated pseudo-ATTs across 500 random permutations. Vertical red dashed lines denote the actual overall summary of aggregated group-time average treatment effects reported in Table 5. Empirical *P* values are obtained from the permutation tests under the null hypothesis of no effect.

Figure 9 demonstrates the alternative event-study estimates obtained by reintroducing the treatment quitters into the full sample (N = 4854). The detailed results are listed in Table S3 in Multimedia Appendix 1. For depressive scores (panel A), with the period immediately preceding initial treatment change (event time *e* = -2) serving as the reference, the post-treatment effects exhibit a statistically significant negative trajectory that increases in magnitude with longer exposure to internet use. This pattern is consistent with our primary findings, indicating that the identified effects of internet adoption on depressive scores are not driven by the exclusion of participants who ever discontinued internet use. For self-rated health (panel B), the effect estimates are bounded tightly around 0 across all periods, aligning with the conclusion of no statistically significant effect. Taken together, these results confirmed that the primary findings in this study, namely, the asymmetric impacts of internet adoption on depressive scores and self-rated health, were robust to behavioral switching in internet use.

**Figure 9.**
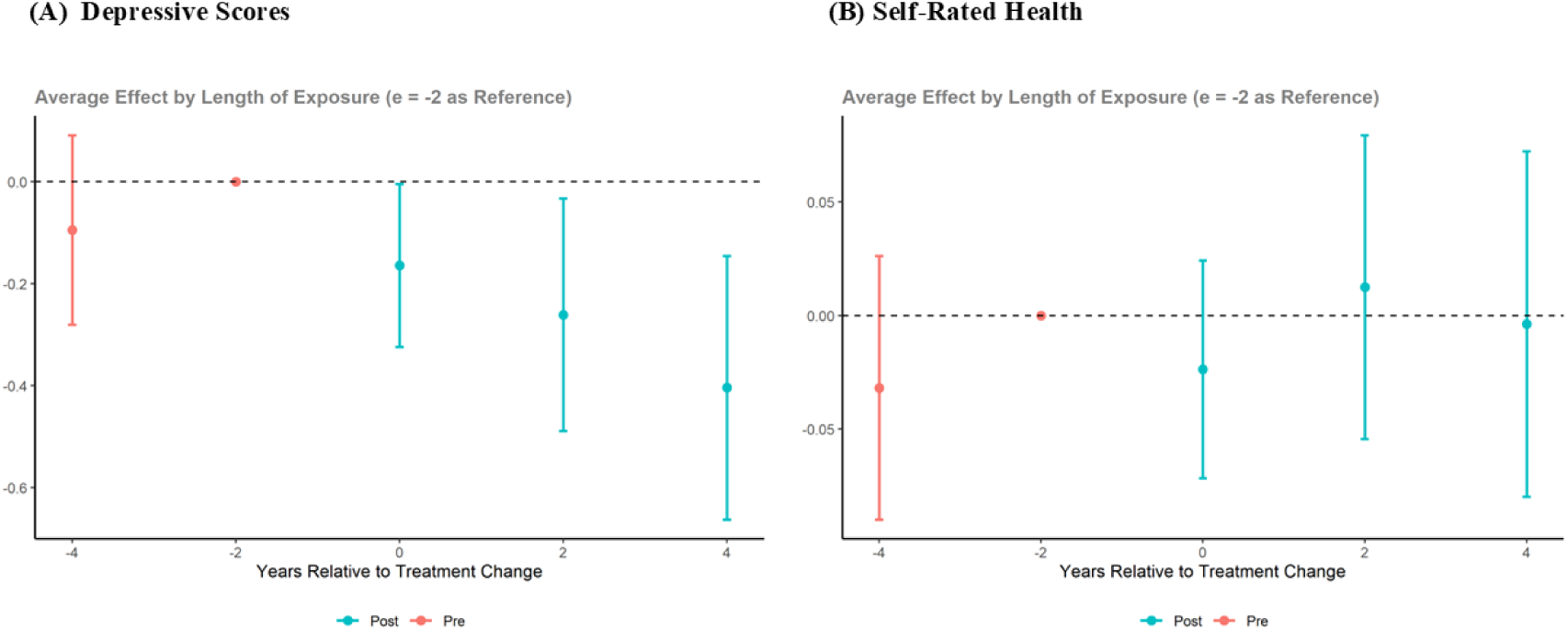
Alternative event-study effects of internet adoption (including treatment quitters). **(A)** Depressive scores; **(B)** Self-rated health. Red dots and error bars represent point estimates and 90% pointwise confidence intervals for pre-treatment periods, whereas blue dots and error bars indicate those for post-treatment periods.

### Mechanism Analysis

Having established the causal effects of internet adoption on both health outcomes, we next explored the underlying mechanisms through which internet adoption operates. To assess the transmission channels comprehensively, we examined 2 categories: individual behavioral variables and household economic variables. We controlled for all pre-treatment covariates for the individual behavioral variables, whereas for each household economic variable we conditioned on the remaining covariates excluding the variable itself. Table 6 reports the conditional parallel trends test results and the corresponding simple weighted average treatment effects of internet adoption. The test results indicate violations of the conditional parallel trends assumption for alcohol consumption (*P* = .070) and per capita family income (*P* = .073) at the 10% significance level. To preserve identification rigor, we focused the mechanism analysis on the 4 channels that satisfied the assumption. Specifically, internet adoption had statistically significant positive effects on all 4 variables, increasing smoking (overall ATT = 0.026, SE = 0.009, *P* < .01), physical exercise frequency (overall ATT = 0.150, SE = 0.049, *P* < .01), media entertainment time (overall ATT = 0.860, SE = 0.424, *P* < .05), and total family expenditure (overall ATT = 0.080, SE = 0.032, *P* < .05).

**Table 6.** Mechanism analysis results.

|  | Conditional parallel trends test, *P* value | Overall ATT (SE) |
|---|---|---|
| **Individual behavioral variables** |  |  |
| Smoking | .370 | 0.026*** (0.009) |
| Alcohol consumption | .070 | 0.012 (0.010) |
| Exercise frequency | .245 | 0.150*** (0.049) |
| Media entertainment | .906 | 0.860** (0.424) |
| **Household economic variables** |  |  |
| Per capita family income | .073 | 0.027 (0.044) |
| Total family expenditure | .175 | 0.080** (0.032) |

Note. Standard errors are presented in the parentheses; \**P* < .10; \*\**P* < .05; \*\*\**P* < .01.

Figure 10 demonstrates the event-study estimates for these variables, illustrating the treatment effect heterogeneity with respect to the length of exposure to continuous internet use. Detailed estimates and standard errors are reported in Table S4 in Multimedia Appendix 1. First, the effect of internet adoption on smoking (panel A) gradually increases in magnitude over time, suggesting that internet use may foster sedentary behavior and associated unhealthy habits. Second, internet adoption is estimated to prompt an immediate increase in physical exercise frequency (panel B), with this positive effect remaining positive across post-treatment periods. This pattern suggests that the broadly accessible online health-promoting content may heighten individual health awareness, potentially translating into sustained physical exercise habits. Third, the effect on media entertainment time (panel C) keeps increasing in magnitude with longer exposure duration. Internet adoption broadens access to digital leisure, with users’ engagement in audio-visual entertainment accumulating as online media becomes increasingly diverse and abundant. Finally, the effect on total family expenditure (panel D) follows a distinct U-shaped trajectory. Upon initial adoption, internet access expands consumption channels, generating an immediate spending stimulus. The effect then temporarily subsides, potentially reflecting a consumption rationalization, before rebounding after 4 years of continuous exposure, likely driven by the established online purchasing habits and diversified digital consumption options.

**Figure 10.**
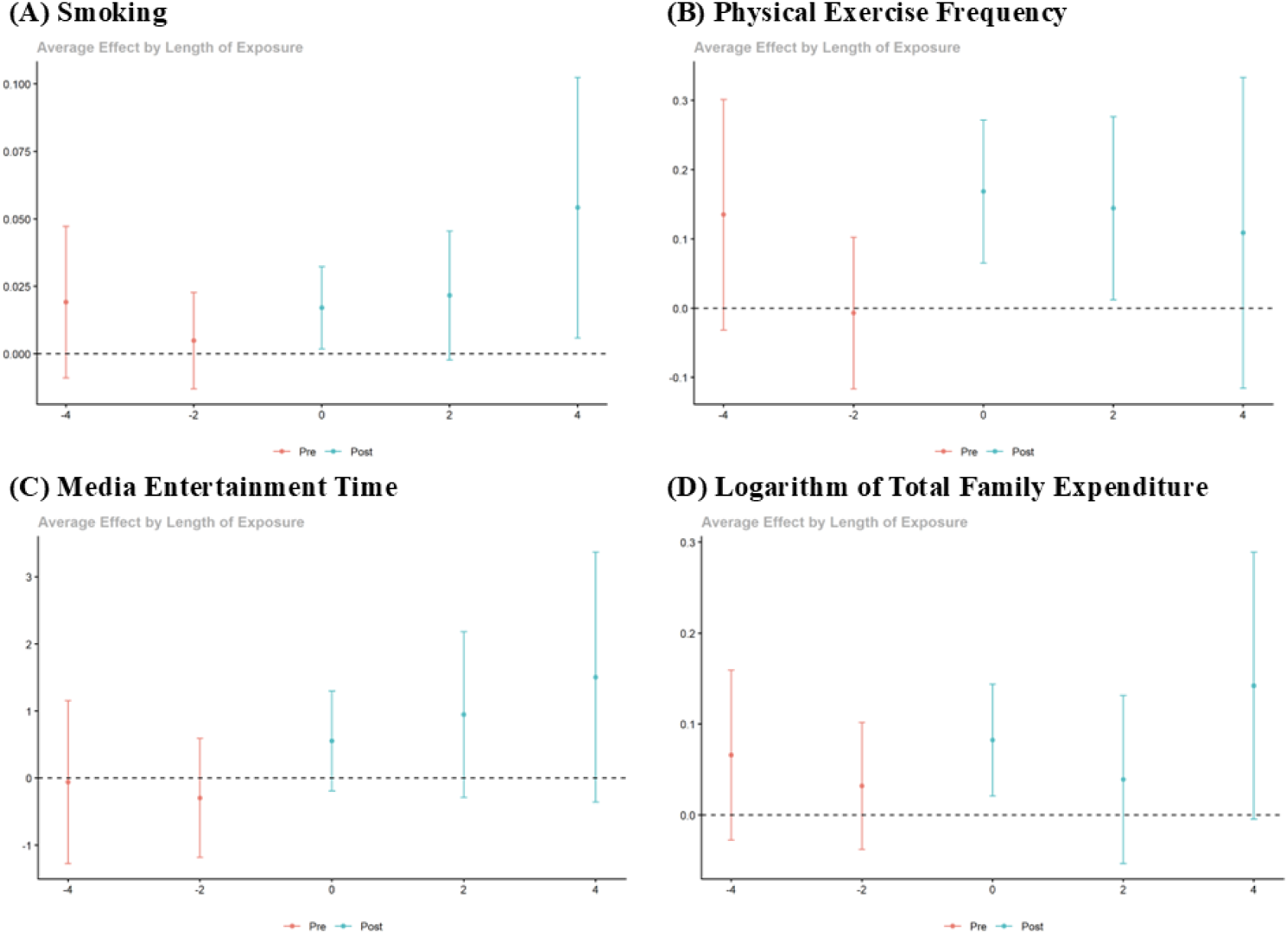
Average treatment effects of internet adoption on individual behavioral and household economic variables based on event-study aggregation. **(A)** Smoking; **(B)** physical exercise frequency; **(C)** media entertainment time; **(D)** logarithm of total family expenditure. Red dots and error bars represent point estimates and 90% simultaneous confidence intervals for pre-treatment periods, whereas blue dots and error bars represent those for post-treatment periods.

Together, these event-study effect trajectories elucidate the mechanisms driving the observed asymmetric health return pattern. On the one hand, psychological well-being benefited through several channels. Grounded in mood management theory [28,29], media entertainment facilitates affective self-regulation, thereby relieving stress and generating positive emotions [30–32]. Expanding engagement in media entertainment offers a sustained pathway for emotional regulation and mood optimization [29,33]. Concurrently, regular physical activity reduces depressive symptoms and alleviates mental health burdens [34–37] by building emotional resilience and self-efficacy [38,39]. Regarding household economics, internet adoption substantially stimulated total household expenditure, consistent with previous studies [40]. Empirical evidence suggests that higher consumption levels positively influence subjective well-being and life satisfaction across age groups [41–43]. Even smoking, despite being a health-compromising behavior, has been documented to provide short-term stress relief [44]. On the other hand, the absence of a statistically significant effect on self-rated health potentially reflects a dynamic behavioral counterbalance [45]. Although regular physical exercise confers multidimensional physical health benefits [46–48], these physiological gains are likely offset by the adverse health consequences of increased smoking and prolonged sedentary behavior [49,50], which accounts for the null effect on self-rated health.

## Discussion

### Principal Findings

Using longitudinal data from 4 waves of the nationwide CFPS spanning 2016-2022, this study examined the causal effects of internet adoption on depressive scores and self-rated health among baseline internet nonusers. To overcome weighting bias and contaminated comparisons inherent in standard TWFE regression under staggered treatment adoption, we employed the DiD with multiple time periods framework, conditioning on a comprehensive set of pre-treatment covariates. Our empirical findings revealed an asymmetric health return pattern, wherein internet adoption produced a cumulative reduction in depressive scores relative to what would have happened in the absence of internet adoption, whereas it had no statistically significant effect on self-rated health. These findings remained robust to participants’ behavioral switching in internet use and were not artifacts of random chance.

Descriptive statistics of the study sample showed a gradual increase in depressive scores from 2016 to 2022, reflecting a macro-level rise in the burden of depression in China documented in the literature [51]. Against this prevailing trend, however, our findings indicated that internet adoption significantly reduced depressive scores relative to the non-adoption counterfactual, in line with existing evidence on the psychological benefits of digital access [5,6,8]. Crucially, the overall average treatment effect of internet adoption was -0.302 on the CES-D 8 scale (simple aggregation), a magnitude comparable to the average per-wave increase in depressive scores across the study period. As exposure duration extended, the estimated reduction intensified, exceeding 0.50 points after 4 years of continuous internet use. Collectively, these effect magnitudes suggest that internet adoption can function as an effective psychological buffer, mitigating secular increases in depressive symptoms.

The mechanism analysis results further identified the behavioral and economic pathways underlying the asymmetric effects on depressive scores and self-rated health. Internet adoption diversifies access to digital leisure, with cumulative engagement in media entertainment facilitating affective self-regulation and stress relief [29,31,32]. Meanwhile, internet adoption increases physical exercise frequency, which may be motivated by greater exposure to online health information and heightened health awareness, offering physiological benefits [47,48] while building emotional resilience to alleviate depressive symptoms [35,36,38]. Furthermore, internet adoption expands consumption channels and stimulates total household expenditure, and higher consumption levels are linked to greater life satisfaction and lower depressive risk [42,43]. Conversely, these health-promoting shifts are accompanied by prolonged sedentary behavior and health-compromising habits such as smoking [49]. Ultimately, these physiological detriments may offset the physical gains conferred by increased physical exercise, accounting for the null overall effect on self-rated health.

### Implications

The empirical findings from this study have important practical implications for maximizing the public health dividends of digital transformation. First, given that internet adoption serves as a psychological buffer against rising population-level depressive symptoms, policies should prioritize the inclusive expansion of digital infrastructure and affordable connectivity to ensure equitable internet access across all socioeconomic strata. Importantly, as baseline internet nonusers and late adopters disproportionately comprise older adults and socioeconomically disadvantaged groups, hardware access alone is insufficient. Policies should advance age-friendly digital adaptations to lower the initial onboarding barriers and help older adults achieve sustained internet use. Second, since digital engagement elicits a double-edged behavioral response, public health strategies should extend beyond simply expanding internet connectivity. Building on the critical roles of media entertainment and physical activity, digital governance should emphasize the curation and promotion of accessible, health-oriented digital content. Enriching the online ecosystem with engaging multimedia resources and structured digital health campaigns can amplify the positive health effects of internet use. Meanwhile, interventions should foster comprehensive digital health literacy to guide internet users toward moderate and constructive online behaviors. Finally, targeted preventive measures are needed to counteract the health-compromising behaviors associated with excessive internet usage [52,53], which can help translate psychological gains into net improvements in overall health status.

### Limitations

Several limitations of this study warrant discussion. First, regarding variable measurement, internet use was defined as a binary indicator to mitigate measurement error and attenuation bias associated with self-reported usage time. However, this binary measure lacks functional granularity as it does not differentiate among specific online activities conducted by internet users. Additionally, both depressive scores and self-rated health were assessed via self-reported scales, which may introduce reporting or social desirability bias compared with objective clinical assessments. Beyond self-report bias, relying solely on these 2 measures fails to capture the multidimensional nature of mental and physical health. Incorporating a broader array of indicators would enable future research to evaluate the diverse health effects of internet adoption more comprehensively. Second, regarding data timing, although the multi-period DiD framework identified dynamic causal effects across a 6-year window (2016-2022), data constraints and survey item changes across CFPS waves precluded extending the observation window or examining additional potential mechanisms. Moreover, because CFPS collected data biennially at discrete time points, the recorded measurements represent snapshots that cannot capture detailed intra-wave fluctuations and may be sensitive to the conditions during the interview, limiting their representativeness. Third, methodologically, although conditioning on a comprehensive set of baseline covariates supports the conditional parallel trends assumption, the influence of unmeasured time-varying confounders cannot be entirely ruled out. Furthermore, the adopted framework entails trade-offs in subgroup heterogeneity analysis. Unlike conventional linear models, this method precludes the direct inclusion of treatment-covariate interaction terms for identifying how specific covariates moderate the treatment effect, and stratifying the sample into subgroups would require the conditional parallel trends assumption to hold independently within each stratum. Consequently, exploring treatment effect heterogeneity for depressive scores and self-rated health across demographic and socioeconomic subgroups requires future investigation.

In terms of research design, this study employed a quasi-experimental approach using secondary observational data. Although the sample was nationally representative, future research would benefit from integrating prospective experimental designs, detailed digital behavioral data, or randomized controlled trials targeting specific online behaviors. Finally, since these findings are situated within a specific timeframe characterized by China’s unique socio-institutional context and rapid digital expansion, caution should be exercised when generalizing the conclusions to other time periods or to countries with differing demographic structures, cultural norms, and health systems.

## Conclusions

This study investigated the population-level causal effects of internet adoption on depressive scores and self-rated health among baseline internet nonusers in China across 4 waves of CFPS data from 2016 to 2022, using a DiD framework with multiple time periods. Over this 6-year window, internet adoption produced a distinct asymmetric health return pattern, characterized by cumulative reduction in depressive scores relative to the non-adoption counterfactual alongside no statistically significant effect on self-rated health. These findings remained robust to participants’ behavioral switching in internet use and were not artifacts of random chance. Mechanism analyses indicated that internet adoption likely alleviated depressive symptoms through expanded media entertainment time, increased physical exercise frequency, and higher household expenditure, whereas the physiological gains of physical exercise were counterbalanced by screen-induced sedentary behaviors and health-compromising behaviors such as smoking. These findings provide critical insights into maximizing the public health dividends of digital transformation across the population. Tailored public health and digital governance strategies are needed to expand inclusive digital infrastructure and age-friendly adaptations, foster digital health literacy, and promote health-oriented digital content. Meanwhile, targeted preventive measures should be established to counteract the physical detriments of excessive internet usage. Future research should complement these macro-level observational findings by integrating granular digital behavioral data or conducting prospective randomized controlled trials. Subsequent studies may focus on exploring demographic and socioeconomic subgroup heterogeneity and evaluating the generalizability of these findings across broader socio-institutional contexts.

## Acknowledgements

We express our gratitude to the Institute of Social Science Survey at Peking University for conducting the China Family Panel Studies project and publishing the data. We thank all survey participants for their involvement.

## Data Availability

The primary data used in this study are available from the China Family Panel Studies Data Platform [54]. The processed datasets generated during the study and the analytical code are available from the authors upon reasonable request.

## Authors’ Contributions

Conceptualization and methodology: YY and YDZ

Data curation and analysis: YY

Writing—original draft: YY

Writing—review and editing: YY and YDZ

## Abbreviations

ATT: Average Treatment Effect for the Treated
CES-D 8: Center for Epidemiologic Studies Depression Scale Short Form
CFPS: China Family Panel Studies
DiD: Difference-in-Differences
TWFE: Two-Way Fixed Effects

## Multimedia Appendix 1

Tables S1, S2, S3, and S4.

