## Supplementary material for "Effects of Internet Adoption on Depressive Scores and Self-Rated Health in China: Multi-Period Difference-in-Differences Analysis": Tables S1, S2, S3, and S4.

**Table S1. Group-time average treatment effects of internet adoption under the conditional specification.**

| Cohort *g* | Time *t* | Depressive scores | Self-rated health |
|---|---|---|---|
| | | ATT (*g*, *t*) (SE) | ATT (*g*, *t*) (SE) |
| 2018 | 2018 | -0.238 (0.263) | 0.017 (0.084) |
| 2018 | 2020 | -0.175 (0.312) | 0.161* (0.089) |
| 2018 | 2022 | -0.568** (0.278) | 0.040 (0.096) |
| 2020 | 2018 | 0.240 (0.229) | -0.011 (0.071) |
| 2020 | 2020 | -0.387* (0.226) | 0.022 (0.072) |
| 2020 | 2022 | -0.436** (0.219) | 0.013 (0.078) |
| 2022 | 2018 | -0.053 (0.204) | -0.005 (0.058) |
| 2022 | 2020 | -0.059 (0.193) | 0.023 (0.057) |
| 2022 | 2022 | -0.019 (0.214) | -0.041 (0.057) |

Note. Standard errors are presented in the parentheses; *$P$ < .10; **$P$ < .05; ***$P$ < .01.

**Table S2. Average treatment effect estimates of internet adoption across aggregation dimensions.**

| | Depressive scores, effect (SE) | Self-rated health, effect (SE) |
|---|---|---|
| **Group-specific effects** | | |
| Cohort 2018 | -0.327 (0.252) | 0.073 (0.072) |
| Cohort 2020 | -0.411** (0.206) | 0.017 (0.063) |
| Cohort 2022 | -0.019 (0.203) | -0.041 (0.056) |
| **Event-study effects (event time *e* = *t* - *g*)** | | |
| Pre-treatment (*e* = -4) | -0.053 (0.204) | -0.005 (0.058) |
| Pre-treatment (*e* = -2) | 0.080 (0.122) | 0.007 (0.041) |
| Post-treatment (*e* = 0) | -0.209* (0.112) | -0.001 (0.035) |
| Post-treatment (*e* = 2) | -0.286 (0.223) | 0.098 (0.063) |
| Post-treatment (*e* = 4) | -0.568* (0.308) | 0.040 (0.086) |
| **Calendar-time effects** | | |
| 2018 | -0.238 (0.263) | 0.017 (0.084) |
| 2020 | -0.266 (0.218) | 0.102 (0.069) |
| 2022 | -0.350** (0.159) | 0.006 (0.053) |

Note. Standard errors are presented in the parentheses; *$P < .10$; **$P < .05$; ***$P < .01$.

**Table S3. Alternative event-study effect estimates of internet adoption (including treatment quitters).**

| | Depressive scores, effect (SE) | Self-rated health, effect (SE) |
|---|---|---|
| Pre-treatment (*e* = -4) | -0.095 (0.113) | -0.032 (0.035) |
| Pre-treatment (*e* = -2) [ref.] | - | - |
| Post-treatment (*e* = 0) | -0.164* (0.097) | -0.024 (0.029) |
| Post-treatment (*e* = 2) | -0.261* (0.139) | 0.013 (0.041) |
| Post-treatment (*e* = 4) | -0.404** (0.157) | -0.004 (0.046) |
| Average cumulative effect | -0.295* (0.119) | -0.008 (0.036) |

Note. Standard errors are presented in the parentheses; *$P < .10$; **$P < .05$; ***$P < .01$.

**Table S4. Event-study effects of internet adoption on individual behavioral and household economic variables.**

| | Smoking | Exercise frequency | Media entertainment | Household expenditure |
|---|---|---|---|---|
| | Effect (SE) | Effect (SE) | Effect (SE) | Effect (SE) |
| Pre-treatment (*e* = -4) | 0.019<br>(0.012) | 0.135<br>(0.075) | -0.060<br>(0.546) | -0.066<br>(0.041) |
| Pre-treatment (*e* = -2) | 0.005<br>(0.008) | -0.007<br>(0.049) | -0.295<br>(0.399) | 0.032<br>(0.030) |
| Post-treatment (*e* = 0) | 0.017**<br>(0.007) | 0.169***<br>(0.046) | 0.553*<br>(0.336) | 0.082***<br>(0.027) |
| Post-treatment (*e* = 2) | 0.022**<br>(0.010) | 0.144**<br>(0.059) | 0.947*<br>(0.556) | 0.039<br>(0.040) |
| Post-treatment (*e* = 4) | 0.054**<br>(0.021) | 0.109<br>(0.101) | 1.505*<br>(0.839) | 0.142**<br>(0.064) |
| Overall dynamic ATT | 0.031***<br>(0.011) | 0.141**<br>(0.057) | 1.002**<br>(0.492) | 0.088**<br>(0.036) |

Note. Standard errors are presented in the parentheses; *$P < .10$; **$P < .05$; ***$P < .01$.
